# Obesity, walking pace and risk of severe COVID-19: Analysis of UK Biobank

**DOI:** 10.1101/2020.07.10.20150003

**Authors:** Thomas Yates, Cameron Razieh, Francesco Zaccardi, Samuel Seidu, Melanie J Davies, Kamlesh Khunti

**Author notes:** **Corresponding author:** Thomas Yates. Prof. Kamlesh Khunti is member of the independent SAGE group. No other conflicts have been declared.

## Abstract

Obesity is an emerging risk factor for coronavirus disease-2019 (COVID-19). Simple measures of physical fitness, such as self-reported walking pace, could also be important risk factors, but have not been well documented. This analysis includes 414,201 UK Biobank participants with complete covariate and linked COVID-19 data. We analysed the risk of severe (in-hospital) COVID-19 across categories of obesity status and walking pace. As of June 20^th^ 2020 there were 972 cases of severe COVID-19 that had occurred within the cohort. Compared to normal weight individuals, the adjusted odds ratio (OR) for severe COVID-9 in those with obesity was 1.49 (1.24, 1.78). Compared to those with a brisk walking pace, the OR in slow walkers was 1.84 (1.49, 2.27). Slow walkers had the highest risk of severe COVID-19 regardless of obesity status. For example, compared to normal weight brisk walkers, the odds of severe COVID-19 in obese brisk walkers was 1.39 (0.99, 1.98), whereas the odds in normal weight slow walkers was 2.48 (1.56, 3.93). Self-reported walking pace, a simple measure of functional fitness, appears to be a risk factor for severe COVID-19 that is independent of obesity. This may help inform simple pragmatic public health risk stratification and preventative strategies.

## Introduction

The severe acute respiratory syndrome coronavirus 2 (SARS-CoV-2), which causes coronavirus disease-2019 (COVID-19), has devastated global economies and put unprecedented strain on health care services and communities. The mortality and economic burden caused by the virus have led to global research efforts to identify people at greatest risk of developing severe illness. Current evidence suggests that subjects with cardiometabolic diseases are at particularly high risk of severe SARS-CoV-2 infection and resulting complications (1). There is also emerging evidence that obesity may be one of the key unifying factors underpinning this risk, as excess ectopic fat accumulation is associated with a higher chronic sub-clinical inflammation, functional immunologic deficit and a pro-thrombotic state potentially explaining the higher rates of disseminated intravascular coagulation and thromboembolism in severe COVID-19 patients (2). However, less is known about the importance of other global markers of physical health. Self-reported walking pace, a measure of functional fitness, has been shown to be a strong predictor of mortality (3,4). Subjects with a self-reported slow walking pace have 2 to 4 times the risk of cardiovascular mortality compared to brisk walkers, whilst also being estimated to die up to 20 years earlier (3,5). Indeed, self-reported walking pace has been shown to be a stronger predictor of cardiovascular mortality than other measures of physical activity or function (6,7). These strong associations with cardiovascular health are thought to reflect the fact that walking pace is a powerful marker of cardiopulmonary function and acts as a global measure of whole-body physical fitness, reserve and resilience (6). Importantly, the strength of association between walking pace and health outcomes has been shown to vary by obesity status (3,5). As COVID-19 primarily affects the cardiopulmonary system, self-reported walking pace is a key target for investigation with important implications for future COVID-19 research and public health.

The aim of this brief report is to examine the relative association of BMI and walking pace with the risk of severe COVID-19.

### Methods

For this analysis, we used UK Biobank (https://www.ukbiobank.ac.uk/), a large prospective cohort of middle-aged adults designed to support health research (8). Between March 2006 and July 2010, 502,543 individuals living within 25 miles of one of the 22 study assessment centres located throughout England, Scotland and Wales were recruited and provided comprehensive data on a broad range of demographic, clinical, lifestyle, and social factors. UK Biobank data is linked to Public Health England’s Second Generation Surveillance System for all national SARS-CoV-2 laboratory test data collected throughout England (9). Data provided included specimen date and origin (hospital inpatient vs other). Data were available for the period 16^th^ March 2020 to June 20^th^ 2020. We restricted the analysis to those from English centres and those that were alive as of 16^th^ March and thus covered by the linkage system. We have classified a positive test result from an in-hospital setting as defining severe COVID-19, consistent with the detailed linkage information provided for this dataset (9).

BMI and walking pace were collected at baseline assessment. BMI was categorised as normal weight (<24.9 kg/m^2^), overweight (25-29.9 kg/m^2^), and obese (≥30kg/m^2^). Habitual walking pace was self-reported as slow (<3mph), steady/average (3-4 mph), or brisk (>4 mph) and has been shown to be associated with cardiorespiratory fitness (3). Mutually exclusive categories of walking pace and obesity status were created to investigate the odds of severe COVID-19 across possible combinations of these factors.

Medical history, including prevalent illnesses, were assessed by interview during baseline visit; ethnicity was assessed by self-report and social deprivation by the Townsend Index (a composite measure of deprivation based on unemployment, non-car ownership, non-home ownership, and household overcrowding; negative values represent less deprivation). Age was measured at March 16^th^ 2020, the first available day for linkage used in this analysis.

We undertook logistic regression to analyse the association of obesity status, walking pace and their combination with the risk of severe COVID-19 as of June 20^th^th 2020. Models were adjusted for the covariates age, sex, ethnicity, social deprivation, number of reported illnesses per person, and the follow-up time from baseline data collection. Data are reported as odds ratios (OR) (95% CI). CIs that do not cross one (reference) were considered significant at p < 0.05.

## Results

There were 414,201 individuals that were alive from English centres with complete covariate data, and thus eligible for coverage by the data linkage system. In total, there were 972 cases of severe COVID-19 that occurred within this cohort. The characteristics of the included participants, stratified by BMI category, are reported in **Table 1**.

**Table 1:** Participant characteristics stratified by obesity and walking pace status.

|  |  | Normal weight<br>(n = 139,185) |  |  | Overweigh<br>(n = 176,421) |  |  | Obese<br>(n = 98,595) |  |  |
| --- | --- | --- | --- | --- | --- | --- | --- | --- | --- | --- |
| Outcome |  | N | % |  | N | % |  | N | % |  |
| Severe COVID-19 cases |  | 220 | 0.2 |  | 417 | 0.2 |  | 335 | 0.3 |  |
| Categorical factors |  | N | % |  | N | % |  | N | % |  |
| Sex | Female | 91846 | 66 |  | 84010 | 48 |  | 52794 | 54 |  |
|  | Male | 47339 | 34 |  | 92411 | 52 |  | 45801 | 46 |  |
| Ethnicity | White European | 131964 | 95 |  | 166459 | 94 |  | 92138 | 93 |  |
|  | Other | 7221 | 5 |  | 9962 | 6 |  | 6457 | 7 |  |
| Walking Pace | Brisk | 75013 | 54 |  | 69107 | 39 |  | 15822 | 21 |  |
|  | Steady/average | 59717 | 43 |  | 97409 | 55 |  | 61944 | 63 |  |
|  | Slow | 4455 | 3 |  | 9905 | 6 |  | 15822 | 16 |  |
| Continuous factors |  | Median | 25<sup>th</sup> | 75<sup>th</sup> | Median | 25<sup>th</sup> | 75<sup>th</sup> | Median | 25<sup>th</sup> | 75<sup>th</sup> |
| Age at censoring date (years) |  | 67 | 60 | 73 | 69 | 61 | 74 | 69 | 62 | 74 |
| Social deprivation (Townsend score) |  | -2.73 | -3.31 | .16 | -2.28 | -3.70 | .20 | -1.74 | -3.39 | 1.23 |
| Diseases or illnesses (n per person) |  | 1 | 0 | 2 | 1 | 1 | 3 | 2 | 1 | 4 |

25<sup>th</sup> and 75<sup>th</sup> indicate 25<sup>th</sup> and 75<sup>th</sup> percentile, respectively

Both BMI and walking pace were independently associated with the risk of severe COVID-19. Compared to normal weight individuals, the adjusted OR of severe COVID-9 in overweight and obese individuals was 1.25 (1.06, 1.48) and 1.49 (1.24, 1.78), respectively. Compared to those with a brisk walking pace, the OR of severe COVID-19 in steady/average and slow walkers was 1.15 (0.99, 1.33) and 1.84 (1.49, 2.27), respectively. The odds of severe COVID-19 across possible combinations of obesity status and walking pace are shown in **Figure 1**. Compared to normal weight brisk walkers, obese steady/average and obese slow walkers had a higher odds of severe COVID-19, but not obese brisk walkers (**Figure 1**). However, compared to normal weight brisk walkers, the odds of severe COVID-19 in slow walkers was over 2 time greater across all categories of obesity status. For example, compared to normal weight brisk walkers, the odds of severe COVID-19 in obese brisk walkers was 1.39 (0.99, 1.98), whereas the odds in normal weight slow walkers was 2.48 (1.56, 3.93).

**Figure 1:**
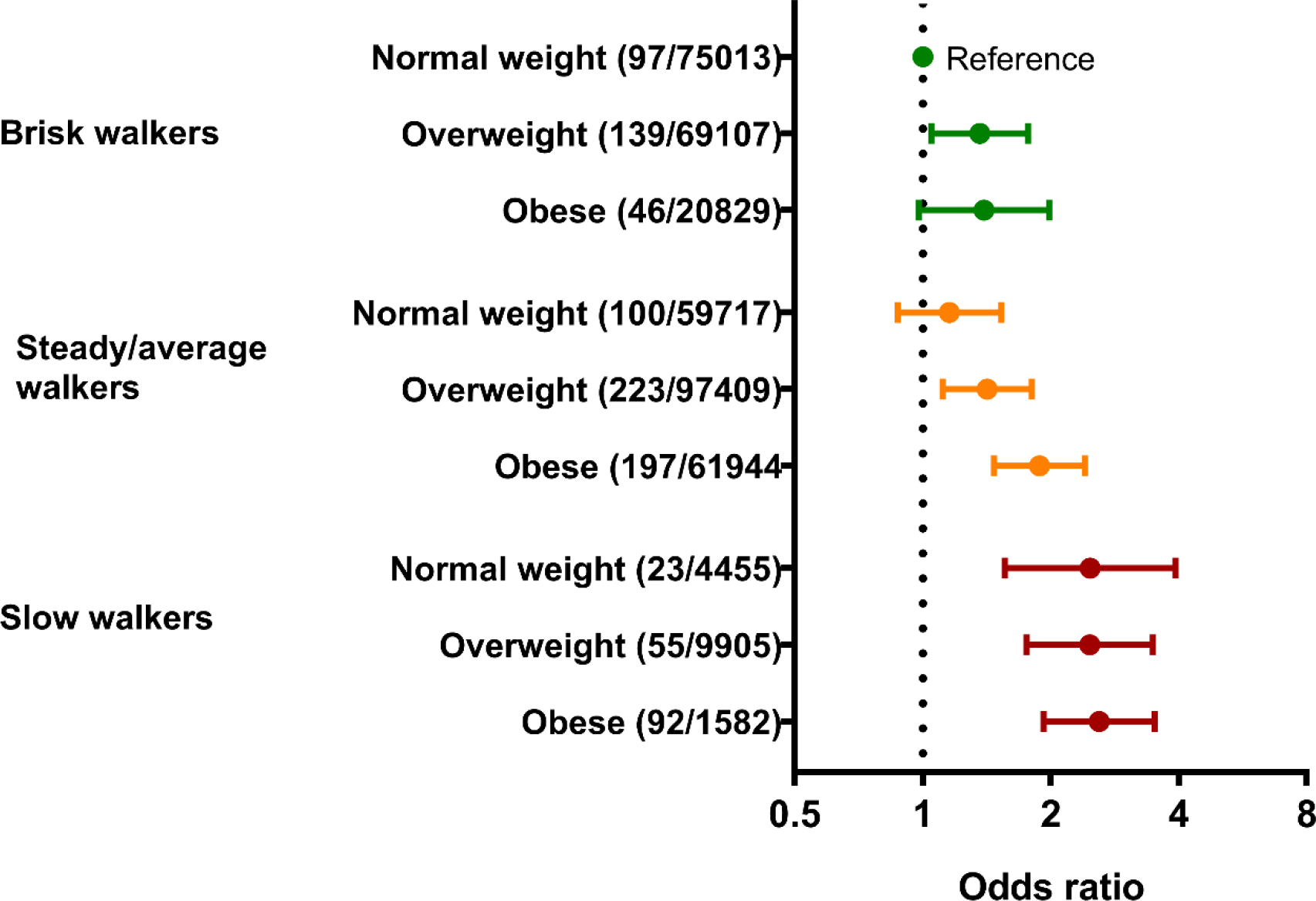
Association of combined obesity and walking pace categories with severe COVID-19.

Data as OR (95 % CI). Numbers display (cases of severe COVID-19/total number in category). Adjusted for age, sex, ethnicty, number of illnesses, social deprivation, and follow-up time from baseline assessment.

## Discussion

Both obesity and walking pace are independently associated with the population level risk of severe COVID-19 in UK Biobank. However, slow walkers had the highest risk of severe COVID-19 regardless of their obesity status, with normal weight, overweight or obese slow walkers all having over twice the risk of severe COVID-19 compared to normal weight brisk walkers. These findings provide further novel evidence that traditional risk factors for cardio-metabolic disease also act as risk factors for COVID-19.

Large routine database studies have recently reported the association of obesity with COVID-19 outcomes (10), with mechanisms hypothesised to include restricted pulmonary function and chronic inflammation (2), with adipose tissue further suggested as an important vial reservoir (11). However, routine clinical databases do not have data on measures of physical function or fitness. Self-reported walking pace has consistently shown to be a powerful independent predictor of cardiovascular and all-cause mortality, including in UK Biobank (3,5-7). This analysis suggests that these previous findings may extend to COVID-19. Walking pace is a complex functional activity, with many factors influencing pace, such as motor control, musculoskeletal health, cardiorespiratory fitness, habitual activity levels, cognition, mental health, and environment factors (7). These factors together may act to give individuals greater resilience to SARS-CoV-2 infection and therefore a lower risk of severe disease.

Previous analysis of walking pace within UK biobank has reported that the risk of cardiovascular mortality or early death may be particular high in those with a combined low BMI and slow walking pace (3, 5), which was hypothesised to result from the greater risk of frailty, under nutrition and poor physical resilience within lean slow walkers compared to obese slow walkers (3, 5). These underlying factors may also explain the findings from the current observations, where slow walkers had a high risk of severe COVID-19 across all categories of obesity status.

The key strength of this study is the large contemporary population with linked COVID-19 data with access to measures of both BMI and walking pace. This study has several important limitations. As testing in the UK has not been universal, it is not possible determine who in the population has been exposed to SARS-CoV-2; linkage will therefore only highlight those that have developed severe disease. Therefore the findings should be interpreted as the overall risk to date in the UK Biobank cohort of being exposed to SARS-CoV-2 and subsequently developing severe COVID-19. Nevertheless, the risk of severe COVID-19 with obesity reported in this study is within the range of early findings reported in other studies (10, 12, 13). As this is an evolving pandemic, data should be interpreted as relating to the first wave of the pandemic in England. Lastly, the risk factors included in this study were measured before the pandemic and therefore do not necessarily reflect participants current health status. However, both BMI and measures of physical fitness have been shown to be stable over time in adults, particularly over relatively short time frames such as a decade (14, 15). The current findings therefore should be interpreted as highlighting the potential importance of simple measures of physical fitness, such as self-reported waking pace, in addition to BMI as potential risk factors for severe COVID-19.

In conclusion, as countries start to ease governmental restrictions to stop the spread of SARS-CoV-2 infection, identifying individuals at greatest risk of developing severe disease is crucial. This study highlights BMI and walking pace as potential risk factors for severe COVID-19, slow walkers in particular having the greatest risk. Ongoing public health and research surveillance studies should therefore consider incorporating simple measures of physical fitness in addition to BMI as potential risk factors that have important public health implications.

## Data Availability

Data are available via UK Biobank

https://www.ukbiobank.ac.uk/

## Acknowledgments

Data were analysed using UK Biobank application number 36371

## Author Contributions

**Concept and design:** C Razieh, T Yates, K Khunti, F Zaccardi

**Acquisition, analysis, or interpretation of data:** All authors

**Drafting of the manuscript:** T Yates

**Critical revision of the manuscript for important intellectual content**: All authors.

**Statistical analysis:** T Yates

**Statistical support**: F Zaccardi

